# Pediatricians’ Attitudes and Knowledge of RSV Immunization Products

**DOI:** 10.1101/2025.05.22.25328182

**Authors:** Costanza Di Chiara, Pierre-Philippe Piché-Renaud, Vera Rigamonti, Anna Cantarutti, Cristina Calvo, Irene Rivero Calle, Antoni Soriano-Arandes, Carlo Giaquinto, Luigi Cantarutti, Daniele Donà, Andrea Lo Vecchio, Adamos Hadjipanayis, Zoi Dorothea Pana, Ioannis Kopsidas, Adria Rose, Alfredo Tagarro, Shaun K Morris, the Penta ID Respiratory Infection working group

**Author notes:** Contributed as co-first authors (Costanza Di Chiara, Pierre-Philippe Piché-Renaud). ***Address correspondence to*:** *Dr. Shaun K. Morris,* MD, Division of Infectious Diseases, Hospital for Sick Children, 555 University Avenue, Toronto, ON, M5G1X8, Canada.

## Abstract

**Purpose:** Nirsevimab and the maternal vaccination in pregnancy are newly introduced prevention strategies against respiratory syncytial virus (RSV). This study evaluated pediatricians’ knowledge, attitudes, and barriers to RSV immunization in Italy and Cyprus/Greece, where nirsevimab had not yet been implemented, and in Spain, where it was introduced in 2023/2024.

**Methods:** A survey was distributed to pediatricians across Spain, Italy, and Cyprus/Greece in July-August/2024. Occupational characteristics, and knowledge, attitudes and practices toward palivizumab, nirsevimab, and the maternal vaccine were collected. Of 578 respondents (response rate=19.3%), 50% were from Spain, 40% from Italy, and 10% from Cyprus/Greece. As 90% of responses came from Spain and Italy, the main analysis focused on these two countries. Descriptive and univariate analyses (Chi-square or Fisher’s exact tests) were conducted.

**Results:** Respondents included pediatricians from community (51.7%), tertiary (30.4%), and primary/secondary care (17.9%) settings. Pediatricians were generally aware of RSV risk factors, without differences across countries. However, 245 (55.9%) and 375 (86.0%) participants reported average-to-low knowledge of nirsevimab and the maternal vaccine, respectively, with differences across countries (p=0.002). Despite limited knowledge, 311 (85.9%) pediatricians were willing to administer nirsevimab, and 319 (92.2%) supported its use for all infants. Nirsevimab administration’s barriers, including unfamiliarity and logistical challenges, were cited by 69 (20.0%) pediatricians. Regarding immunization preferences, 165 (47.4%) pediatricians were very likely to support the maternal vaccine over nirsevimab.

**Conclusions:** While pediatricians support RSV immunization, knowledge gaps and logistical barriers may hinder early-stage implementation. Optimizing education on RSV immunization will be crucial to improve uptake and prevention strategies.

**What’s Known on This Subject:** There is limited evidence regarding pediatricians’ attitudes and knowledge of nirsevimab and maternal RSV vaccination during pregnancy.

**What This Study Adds:** This multi-country survey evaluates pediatricians’ knowledge, attitudes, and barriers to RSV immunization in Italy and Cyprus/Greece, where nirsevimab had not yet been implemented, and in Spain, where it was introduced in 2023/2024.

## Background

Respiratory syncytial virus (RSV) is a leading cause of pediatric lower respiratory infections, resulting in significant hospitalizations even among healthy, term infants [1–5]. Historically, palivizumab, a monthly monoclonal antibody, was the only available RSV prophylaxis, but its high cost limited use to high-risk infants [6,7].

In 2023, the European Medicines Agency (EMA) and the U.S. Food and Drug Administration (FDA) approved nirsevimab, a long-acting monoclonal antibody targeting the highly conserved epitope site Ø on the prefusion form of the RSV fusion (F) protein. Administered as a single dose to all infants during their first RSV season, it demonstrated over 70% efficacy in preventing RSV-related lower respiratory tract infections in both preterm and term infants [8,9], with real-world data showing >80% efficacy in reducing hospitalizations and 85% against severe infections [10–18]. Additionally, both the EMA and FDA approved a maternal RSV vaccine targeting the preF protein, administered in late pregnancy to provide passive immunity to infants. However, it showed a lower efficacy (57%) against severe RSV infection in clinical trials [19].

RSV burden and new preventative tools emphasize the importance of widespread immunization, which relies on healthcare providers, particularly pediatricians, to guide caregivers’ decisions [20–24]. Therefore, understanding pediatricians’ knowledge and attitudes toward RSV and its new immunization strategies is essential for effective implementation [25]. This study assessed pediatricians’ awareness, attitudes, knowledge, and practices regarding RSV immunization in Italy, Cyprus, and Greece–countries where nirsevimab had not yet been adopted–, and in Spain, where it was implemented during the 2023-2024 season. It also compared these factors between countries with and without access to the product.

## Methods

### Study design, setting, and data collection

A multi-language, self-administered, cross-sectional survey was conducted among pediatricians in Spain, Italy, Cyprus, and Greece between July 20 and August 24, 2024. The survey was electronically distributed to 2,000 members of the Spanish Pediatric Association (AEP) (including 346 members of the Spanish Society of Pediatric Infectious Diseases [SEIP]), 180 members of the Italian Society of Pediatric Infectious Diseases, 120 members of the Italian independent network of family pediatricians (Pedianet, Società Servizi Telematici Srl), 250 members of the Cypriot Pediatric Society, and 100 members of the Hellenic Society of Pediatric Infectious Diseases. Each society sent the survey via secure email listservs, with an initial invitation followed by four weekly reminders. The questionnaire was available in Spanish, Italian, Greek, and English.

### Survey instrument

The survey instrument consisted of binary, multiple choice, and Likert scale questions. Developed by the coordinating team at the Hospital for Sick Children, the questionnaire was pilot-tested by general and infectious disease pediatricians to improve clarity. Additionally, all European researchers within the team reviewed the questionnaire to ensure its relevance and appropriateness for their local contexts. Specifically, because nirsevimab was adopted in Spain at the start of the 2023-2024 season, questions regarding palivizumab were asked in the present tense for Italy, Cyprus, and Greece, and in the past tense for Spain, while questions about nirsevimab followed the opposite approach.

At the time of the survey distribution, national recommendations on infant RSV immunization were available only in Spain. The Advisory Committee on Vaccines (CAV) of the Spanish Pediatric Association (AEP) had published its 2024 immunization guidelines, recommending the routine administration of a single dose of nirsevimab for all infants under 6 months of age and an annual dose for high-risk children under 2 years [26]. Additionally, the 2024 CAV-AEP immunization calendar advised administering a dose of maternal RSV vaccination between weeks 24 and 36 of gestation, preferably between weeks 32 and 36, as part of a public health strategy if indicated [26]. Spain’s Ministry of Health prioritized nirsevimab over maternal RSV vaccination [27]. In contrast, in Italy, only a position paper on maternal RSV vaccination during pregnancy had been published by Italian societies of gynecologists and obstetricians [28], while in Greece and Cyprus, no national recommendations on either nirsevimab or maternal RSV vaccination were available at the time of the survey.

The questionnaire included four main sections: 1) pediatricians’ occupational and demographic information, 2) knowledge and perceptions about RSV infection and prevention products, 3) attitude and practices toward palivizumab and nirsevimab, and 4) perspectives on RSV maternal vaccination. The full survey is detailed in Supplementary Materials (**Online Resource 1**).

### Statistical analysis

Descriptive statistics summarized participants’ RSV knowledge, attitudes, and practices. Frequencies and percentages were calculated for each question. It was not required for all questions to be answered by the respondents in order to submit the survey. Incomplete surveys were retained in the analysis, with missing responses being excluded from question-specific calculations of frequencies and percentages.

Specific scores were developed to assess variations in RSV knowledge among pediatricians’ subgroups. These scores were constructed using a core set of key variables and designed to assess two dimensions: (1) knowledge of RSV risk factors, (2) knowledge of RSV prevention products among pediatricians. Each score was calculated by aggregating the key selected variables, normalized, and subsequently categorized into three levels (low, medium, and high). Details on score construction, including the key variables and methodology used, are provided in the supplemental material (**Online Resource 2**). Comparisons of knowledge levels between participant subgroups (country, subspecialty, years of practice, primary practice setting, experience managing RSV cases, and their center’s involvement in vaccination administration) was performed using Chi-square test. Scores were computed exclusively for participants who answered all the key questions necessary for their calculation (hereafter referred to as “having completed the survey”). All statistical analyses were performed using Microsoft Excel software version 2409 and SAS software version 9.4 (SAS Institute, Inc., Cary, NC, USA). Significance was set at P < 0.05.

### Ethics

As part of a broader research initiative conducted in Canada, this study was submitted to and approved by the Hospital for Sick Children’s Research Ethics Board (REB # 1000081077), with protocol acknowledgment in Cyprus and Greece. Due to anonymity, ethics approval wasn’t needed in Spain and Italy. Participation was voluntary, with implied consent upon survey completion. Surveys were completed using the Research Electronic Data Capture (REDCap) software.

## Results

### Participants’ characteristics

After excluding surveys that were only opened (N=56) or only contained country-related responses (country of training and country of practice) (N=69), 578 participants were included (response rate=19.3%). Spain contributed 290 (50%) participants (response rate: 14.5%), Italy 228 (40%) (response rate: 76%), Cyprus 43 (7%) (response rate: 17.2%), and Greece 17 (3%) (response rate: 17%) (**Table 1**, **Online Resource 3**).

**Table 1.** Characteristics of respondents (N=578).

| Characteristics, N (%) | Overall<br>(N=578) | Spain<br>(N=290) | Italy<br>(N=228) | Cyprus<br>(N=43) | Greece<br>(N=17) |
| --- | --- | --- | --- | --- | --- |
| **Pediatric training**<br>**(missing=4)** |  |  |  |  |  |
| General pediatrics | 356 (62.0%) | 181 (62.6%) | 128 (56.9%) | 40 (93.0%) | 7 (41.2%) |
| Subspecialty pediatrics | 218 (38.0%) | 108 (37.4%) | 97 (43.1%) | 3 (7.0%) | 10 (58.8%) |
| **Primary practice setting**<br>**(missing=5)** |  |  |  |  |  |
| Urban | 459 (80.1%) | 238 (82.4%) | 172 (75.8%) | 34 (82.9%) | 15 (93.8%) |
| Suburban | 64 (11.2%) | 26 (9.0%) | 35 (15.4%) | 3 (7.3%) | 0 (0.0%) |
| Rural/remote | 50 (8.7%) | 25 (8.7%) | 20 (8.8%) | 4 (9.8%) | 1 (6.3%) |
| **Type of practice site**<br>**(missing=3)** |  |  |  |  |  |
| Community-based<br>pediatricians | 297 (51.7%) | 145 (50.3%) | 130 (57.0%) | 18 (42.9%) | 4 (23.5%) |
| Primary/secondary care<br>hospital (public and private) | 103 (17.9%) | 41 (14.2%) | 41 (18.0%) | 19 (45.2%) | 2 (11.8%) |
| Academic/Tertiary care<br>hospital | 175 (30.4%) | 102 (35.4%) | 57 (25.0%) | 5 (11.9%) | 11 (64.7%) |
| **Years of practice**<br>**(missing=6)** |  |  |  |  |  |
| <= 5 years | 86 (15.0%) | 38 (13.2%) | 46 (20.4%) | 1 (2.3%) | 1 (5.9%) |
| 6-10 years | 64 (11.2%) | 37 (12.9%) | 24 (10.7%) | 0 (0.0%) | 3 (17.6%) |
| 11-25 years | 180 (31.5%) | 112 (39.0%) | 48 (21.3%) | 12 (27.9%) | 8 (47.1%) |
| >25 years | 242 (42.3%) | 100 (34.8%) | 107 (47.6%) | 30 (7.0%) | 5 (29.4%) |
| **Pediatricians' practices in**<br>**managing cases of**<br>**bronchiolitis**<br>**(missing=2)** |  |  |  |  |  |
| Always | 277 (48.1%) | 151 (52.1%) | 104 (45.6%) | 18 (43.9%) | 4 (23.5%) |
| Very often | 253 (43.9%) | 118 (40.7%) | 107 (46.9%) | 18 (43.9%) | 10 (58.8%) |
| Sometimes | 40 (6.9%) | 19 (6.6%) | 14 (6.1%) | 5 (12.2%) | 2 (11.7%) |
| Rarely | 5 (0.9%) | 2 (0.7%) | 2 (0.9%) | 0 (0.0%) | 1 (5.9%) |
| Never | 1 (0.2%) | 0 (0.0%) | 1 (0.4%) | 0 (0.0%) | 0 (0.0%) |
| **Working in a practice which**<br>**provides immunizations**<br>**(missing=4)** |  |  |  |  |  |
| Yes | 496 (86.4%) | 260 (90.3%) | 183 (80.3%) | 39 (95.1%) | 14 (82.4%) |
| No | 78 (13.6%) | 28 (9.7%) | 45 (19.7%) | 2 (4.9%) | 3 (17.6%) |

Among respondents, 356 (62%) were general pediatricians, and 218 (38%) were subspecialists, mainly in infectious diseases (N=60, 27.5%), emergency medicine/general pediatrics (N=31, 14.2%), and neonatology (N=31, 14.2%) (**Online Resource 4**). Most worked in community settings (N=297, 51.7%), while 103 (17.9%) and 175 (30.4%) were in primary/secondary and academic/tertiary hospitals, respectively. Urban/suburban practice was reported by 523 (91.3%). Over ten years of experience was noted in 422 (73.8%). Regarding bronchiolitis management, 277 (48.1%) “always” and 253 (43.9%) “very often” managed cases. Most (N=496, 86.4%) worked in settings providing immunizations (**Table 1**).

Since 90% of respondents were from Spain and Italy, descriptive analyses and comparisons of RSV knowledge and prevention products are limited to these two countries. Data from Cyprus and Greece are reported as supplemental material. Hereafter, “overall” refers to the 518 participants from Spain and Italy. Among them, 212 fully completed the questionnaire. No significant demographic or occupational differences were observed between those who completed the full questionnaire and the broader Spanish and Italian sample (**Online Resource 5**).

### Knowledge of RSV and prevention products

Overall, most participants correctly identified prematurity (<32 gestational weeks: N=427, 96.2%; 33–35 gestational weeks: N=332, 74.8%), congenital heart disease (N=387, 87.2%), chronic lung disease including bronchopulmonary dysplasia (N=383, 86.3%), and Trisomy 21 (N=243, 54.7%) as risk factors for severe RSV disease. Additionally, 253 (57.4%) participants identified healthy children under two years as at the highest risk of severe RSV disease.

Responses by country are shown in **Online Resource 6** and **Online Resource 7**. Subspecialists were more likely to have medium-to-high RSV risk factors knowledge, while general pediatricians had lower scores (p=0.054) (**Table 2**).

**Table 2.** Level of knowledge of risk factors for severe RSV diseases among 212 respondents from Spain and Italy.

|  |  | Level (Score #1) |  |  | p-value* |
| --- | --- | --- | --- | --- | --- |
|  |  | Low<br>(N=34) | Medium<br>(N=93) | High<br>(N=85) |  |
| **Country** | **Spain** | 20 (58.8%) | 50 (53.8%) | 38 (44.7%) | 0.292 |
|  | **Italy** | 14 (41.2%) | 43 (46.2%) | 47 (55.3%) |  |
| **Pediatric Subspecialty** |  |  |  |  | 0.054 |
|  | **Pediatrician Subspecialist** | 13 (38.2%) | 39 (41.9%) | 49 (57.7%) |  |
|  | **General Pediatrician** | 21 (61.8%) | 54 (58.1%) | 36 (42.4%) |  |
| **Years in practice** |  |  |  |  | 0.073 |
|  | **<=10** | 9 (26.5%) | 23 (24.7%) | 34 (40.0%) |  |
|  | **>10** | 25 (73.5%) | 70 (75.3%) | 51 (60.0%) |  |
| **Description of primary practice** |  |  |  |  | 0.063 |
|  | **Primary care&Primary outpatient center** | 18 (52.9%) | 53 (57.00%) | 33 (38.8%) |  |
|  | **Primary/secondary care hospital&Private hospital** | 2 (5.9%) | 12 (12.9%) | 18 (21.2%) |  |
|  | **Academic hospital&Tertiary care hospital** | 14 (41.2%) | 28 (30.1%) | 34 (40.0%) |  |
| **Manage of RSV cases** |  |  |  |  | 0.569 |
|  | **Always&Very often** | 32 (94.1%) | 85 (91.4%) | 81 (95.3%) |  |
|  | **Sometimes&Rarely&Never** | 2 (5.9%) | 8 (8.6%) | 4 (4.7%) |  |
| **Provision of immunizations to children** |  |  |  |  | 0.120 |
|  | **Yes** | 26 (76.5%) | 83 (89.3%) | 76 (89.4%) |  |
|  | **No** | 8 (23.5%) | 10 (10.8%) | 9 (10.6%) |  |

\*Chi-square test was applied to assess the distribution of demographic and occupational characteristics among respondents with low, medium, or high levels of knowledge, treating these levels as independent variables.

Regarding knowledge of RSV prevention products, 178 (40.5%) respondents reported above average-to-very high knowledge of palivizumab, while 245 (55.9%) and 375 (86.0%) reported average-to-low knowledge of nirsevimab and the maternal RSV vaccine, respectively (**Online Resource 8**). Perceived effectiveness of palivizumab was highest for reducing hospital admissions (N=265, 72.0%), ICU admissions (N=297, 81.3%), and mortality (N=296, 83.3%) (**Online Resource 9**).

Knowledge of prevention products was higher among Spanish pediatricians (p=0.002), reflecting greater familiarity with nirsevimab in Spain, where it was already in use, and among subspecialists (p=0.010) and those who work in academic/tertiary care hospitals (p=0.023) (**Table 3**).

**Table 3.**
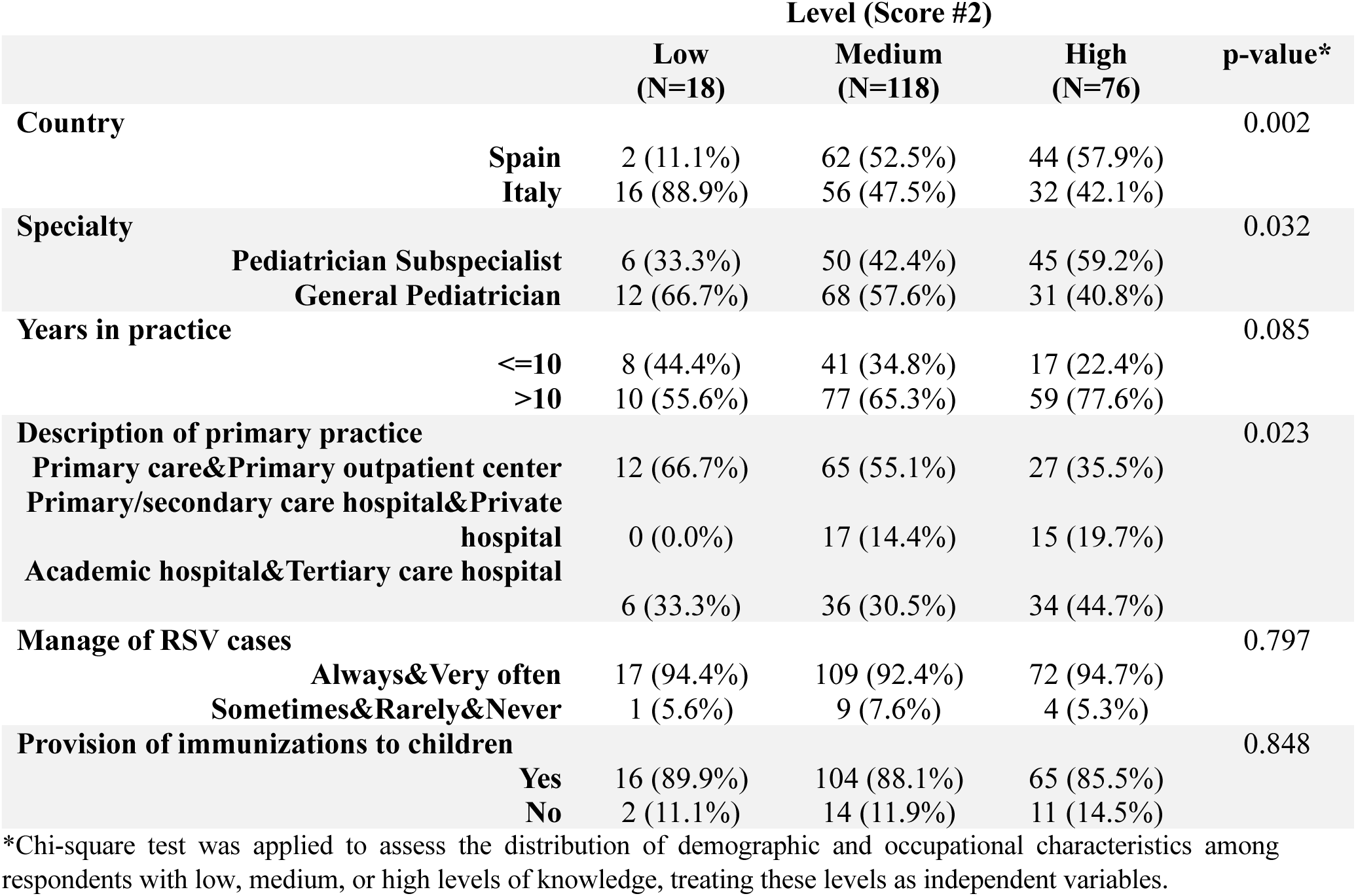
Level of knowledge of RSV prevention products (palivizumab, nirsevimab, and RSV maternal vaccine) among 212 respondents from Spain and Italy.

### Acceptance, attitude, practice, and perception of nirsevimab

Most respondents (N=311, 85.9%) were likely to administer nirsevimab, with 319 (92.2%) indicating they would administer it to all patients, regardless of underlying risk factors (**Table 4, Online Resource 10**). Notably, pediatricians with lower levels of knowledge were less likely to prescribe nirsevimab (**Online Resource 11**).

**Table 4.** Acceptance, attitude, and preference of respondents from Spain and Italy toward nirsevimab administration (N=518).

| **Administration's preference** |  | **N (%)** |
| --- | --- | --- |
| **Likelihood of administering nirsevimab**<br>*(missing=156)* |  |  |
|  | **Very likely** | 311 (85.9%) |
|  | **Likely** | 39 (10.8%) |
|  | **Neutral** | 9 (2.5%) |
|  | **Unlikely** | 1 (0.3%) |
|  | **Very unlikely** | 2 (0.6%) |
| **Target population** |  |  |
|  | **All infants** *(missing=172)* | 319 (92.2%) |
|  | **Only high-risk infants** *(missing=210)* | 51 (16.6%) |
|  | **Only high-risk infants** *(n/51)* |  |
|  | **Preterm birth** | 51 (100.0%) |
|  | **BDP/CLD** | 50 (98.0%) |
|  | **CHD** | 49 (96.1%) |
|  | **Trisomy 21** | 40 (78.4%) |
|  | **Immunocompromised** | 46 (90.2%) |
|  | **Infants lacking access to immediate medical care** | 26 (50.0%) |
|  | **Other** | 2 (3.9%) |
| **Preferred setting**<br>*(missing=154)* |  |  |
|  | **During birth hospitalization** | 292 (80.2%) |
|  | **During the first well-baby visit** | 33 (9.1%) |
|  | **At a specific vaccine clinic** | 17 (4.7%) |
|  | **At primary care centers** | 14 (3.8%) |
|  | **Other** | 8 (2.2%) |
| **Preferred time**<br>*(missing=166)* |  |  |
|  | **Any time of the year** | 118 (33.5%) |
|  | **Seasonal** | 234 (66.5%) |

Regarding preferences, 175 (52.9%) respondents indicated a high likelihood of administering an RSV vaccine to infants over passive immunization, while 28.7% (N=95) reported a moderate likelihood.

Birth hospitalization (N=292, 80.2%) was the most preferred settings for nirsevimab administration. Notably, 118 (33.5%) participants favoured year-round over seasonal administration (**Table 4**).

Perceived barriers to nirsevimab administration were reported by 69 (20.0%) respondents, including concerns about introducing a new product (55.1%), healthcare provider availability (40.6%), and patient access (34.8%) (**Figure 1**). Similarly, 118 (34.3%) respondents mentioned barriers to caregivers’ acceptance of nirsevimab, with the main concerns being caregivers’ perceptions about potential adverse effects (65.3%), lack of awareness about RSV (51.7%), and the administration of a monoclonal antibody to a healthy infant (44.1%) (**Online Resource 12**).

**Figure 1.**
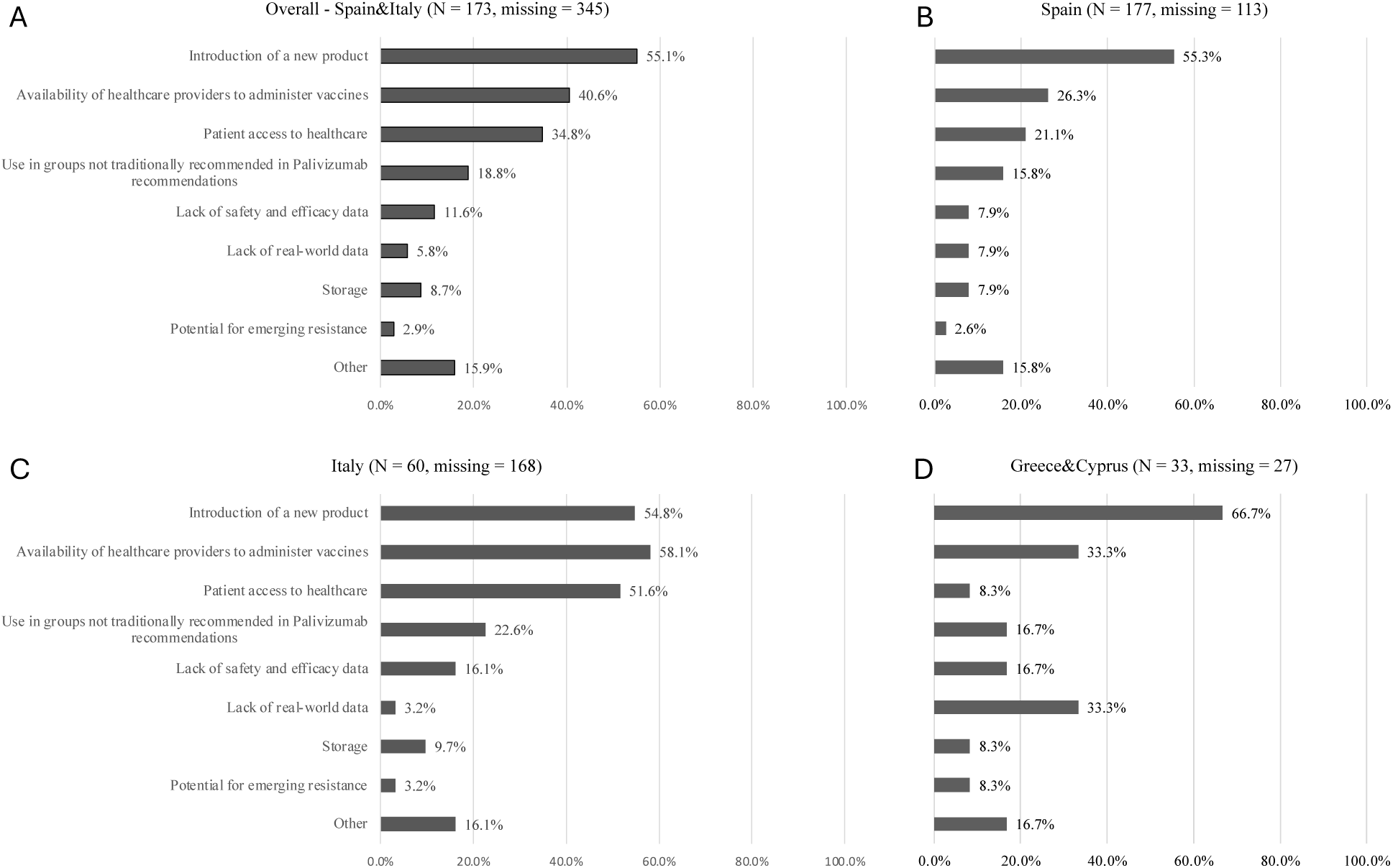
Specific barriers to nirsevimab administration identified among respondents by country.

### Acceptance, attitude, practices, and perception of RSV maternal vaccination

A total of 165 (47.4%) participants were more likely to recommend the RSV maternal vaccine over nirsevimab. However, 125 (38.5%) participants anticipated barriers to RSV maternal vaccine administration, with 60.8%, 58.4%, and 54.4% expressing concerns about introducing a vaccine during pregnancy, administering multiple vaccines during pregnancy, and potential adverse effects, respectively (**Online Resource 13**). Additionally, 113 (32.6%) participants believed that acceptance of the RSV maternal vaccine would be lower or much lower than that of nirsevimab. A total of 145 (45.0%) anticipated barriers to caregivers’ acceptance of RSV maternal vaccine, with the most frequently mentioned concerns being caregivers’ perceptions of potential adverse effects on the infant (81.4%), potential adverse effects on the mother (75.9%), and lack of awareness about RSV (37.2%) (**Online Resource 14**).

While 90 (27.4%) and 112 (34.0%) pediatricians routinely discussed influenza and Tdap vaccination with pregnant women, only 44 (13.4%) consistently addressed COVID-19 vaccination (**Online Resource 15**). Nonetheless, 160 (47.8%) and 134 (40.0%) would “always” or “very often” recommend a multi-respiratory vaccine approach, including RSV, influenza, and COVID-19 vaccines.

## Discussion

New immunization products often face challenges due to uncertainty and lack of confidence. This multi-country survey assessed the attitudes, knowledge, and practices of 578 pediatricians from Spain, Italy, Cyprus, and Greece regarding RSV immunization. To our knowledge, this is the first study to evaluate pediatricians’ perspectives on RSV immunization across multiple European countries, comparing those with and without nirsevimab implementation.

Most participants expressed favorable attitudes toward nirsevimab, consistent with prior studies showing high acceptance among pediatricians in Europe and the USA [29,30]. Notably, 92.2% indicated they would administer it to all infants, aligning with international recommendations [31]. Importantly, the likelihood of prescribing nirsevimab increased with greater knowledge levels, underscoring the critical role of continuous education in shaping prescribers’ attitudes and decision-making. Furthermore, pediatricians with subspecialties demonstrated greater understanding of RSV risk factors, likely due to easier access to educational events [29]. These findings emphasize the need for educational initiatives for both hospital- and community-based pediatricians. As expected, pediatricians in Italy—where nirsevimab was not yet introduced and national recommendation were not published—had lower knowledge of nirsevimab compared to Spanish colleagues but were generally willing to prescribe it, suggesting the importance of educational initiatives for healthcare providers early in the implementation of the RSV programme.

Overall, 20.0% reported anticipated barriers to administration of nirsevimab, such as concerns about introducing a new product, limited healthcare provider availability, and restricted access. These barriers align with those identified by other authors, such as timing, familiarity, cost, reimbursement issues [32], and supply shortages [33]. These findings emphasize the need for strategies like targeted pediatrician training and enhanced logistical support to improve nirsevimab access.

Pediatricians also anticipated caregivers’ barriers to acceptance of nirsevimab, with 65.3% mentioning concerns on caregivers’ perceptions about adverse effects, lack of RSV awareness, and hesitancy to administer monoclonal antibodies to healthy infants. These findings are consistent with previous studies showing that caregivers question nirsevimab’s safety and effectiveness [30], and low parental knowledge of RSV is a barrier to immunization acceptance at birth [34]. Since many caregivers plan vaccination schedules before pregnancy [35], providing early education on RSV during pregnancy could enhance nirsevimab acceptance and uptake. Moreover, a successful immunization program in Spain demonstrated that ensuring easy access to appointments—offered during non-working hours and at diverse locations—can significantly boost immunization rates [36].

Regarding immunization preferences, pediatricians favored administering nirsevimab during birth hospitalization or at the first well-baby visit. While most preferred seasonal administration, 33.5% expressed interest in year-round use. This preference may be particularly relevant for countries with limited primary care resources, as the long-acting nature of nirsevimab could facilitate its integration into routine visits or other immunization services. Such integration has previously been associated with higher vaccine uptake in resource-limited settings [37], as in low- and middle-income countries, common barriers to vaccination include distance to healthcare access points, lack of partner support, and inconvenient timing, which have been reported as key challenges for caregivers [38].

Despite 86.0% of respondents reporting an average-to-low level of knowledge of the RSV maternal vaccine, as well as evidence of its reduced efficacy [19], nearly half of participants would prefer administering the maternal vaccine over nirsevimab, potentially due to earlier introduction of national recommendations of maternal RSV vaccine in pregnancy in Italy. This aligns with previous findings, where around 60% of obstetricians favored preF maternal vaccination [30]. However, 32.6% believed that acceptance of the maternal vaccine would be lower than nirsevimab. About 60% of pediatricians reported regularly discussing maternal vaccinations against respiratory infections with pregnant women, despite evidence that recommending vaccines at each visit and emphasizing infant health is effective for maternal vaccination uptake [39]. This suggests a need for improved maternal vaccination education targeting healthcare providers who care for pregnant women and children.

This study has several strengths, including a large sample size, geographic diversity, and the inclusion of countries both with and without nirsevimab adoption. This design provides a broad evaluation of pediatricians’ attitudes and practices toward RSV prevention. The involvement of leading pediatric societies in each country further facilitated the inclusion of pediatricians who are actively engaged in RSV prevention efforts within their respective regions. However, the study has limitations. The “self-administration” of participants may introduce bias, which could affect the generalizability of the results. Additionally, the sample size was limited, with only 10% of respondents from Cyprus and Greece; as a result, the main analyses focused on data from Spain and Italy, while descriptive data from Cyprus and Greece are reported in the Supplementary Materials. Furthermore, not all participants completed the full survey, potentially affecting the results. The inclusion of uncompleted survey in the analysis is also a limitation. However, since the REDCap function that redirects users to their existing survey when they log in using the same email address was implemented, the risk of duplications and entry errors can be considered limited. Some biases may also arise due to the overrepresentation of specialists in infectious diseases, neonatology, and emergency medicine, who may be more likely to engage in topics related to RSV immunization. However, including neonatologists in the sample is crucial, as they are involved in administering monoclonal antibodies and play a key role in RSV prevention for infants. Moreover, the survey assessed pediatricians’ perceived barriers to the implementation of RSV immunization products, which may have led to a potentially inaccurate representation of caregiver-related barriers due to the lack of direct input from caregivers themselves. Finally, while the survey provides valuable insights, it is limited by its design compared to more robust epidemiological studies and ongoing research will be required as these new products are rolled out in different countries to understand the barriers and facilitators to real world implementation.

## Conclusions

Despite high acceptance of nirsevimab and the RSV maternal vaccine among European pediatricians, knowledge gaps and anticipated barriers to administration remain. Targeted educational initiatives to optimize pediatric practices regarding RSV infant immunization and maternal vaccination may be beneficial. Addressing these gaps could improve implementation and increase immunization rates, protecting vulnerable populations from RSV and other respiratory viruses.

## Abbreviations

RSV: Respiratory Syncytial Virus
EMA: European Medicines Agency
FDA: U.S. Food and Drug Administration
REDCap: Research Electronic Data Capture
SMD: Standardized Mean Difference

## Acknowledgments

The corresponding author acknowledges Drs. Upton Allen, Shelly Bolotin, Malini Dave, Tiffany Fitzpatrick, Ryan Huang, Julia Orkin, and Michelle Science for their contributions to the development of the survey. Drs. Elahe Karimi-Shahrbabak and Andrea Oletto are thanked for their support with the implementation of the survey instrument on REDCap. The corresponding author also extends thanks to Dr. Federica D’Ambrosio for her role in managing the study within the Penta Respiratory Infections Working Group.

The authors acknowledge the collaboration of the following scientific societies: the Spanish Society of Paediatric Infectious Diseases (SEIP), the Spanish Pediatric Association (AEP), the Italian Society of Pediatric Infectious Diseases, the Italian independent network of family pediatricians (Pedianet, Società Servizi Telematici Srl), the Cypriot Pediatric Society, and the Hellenic Society of Pediatric Infectious Diseases. The authors also thank all the pediatricians who participated in the project.

## Statements and Declarations

### Conflict of interest disclosures (includes financial disclosures)

IRC has received speaking fees from MSD, GSK, Sanofi, Moderna and Pfizer; scholarships/research grants from Sanofi Pasteur, MSD, Novartis and Pfizer; consulting fees for Pfizer, MSD, Sanofi; and has participated as a subinvestigator in clinical trials of vaccines from Ablynx, Abbot, Seqirus, Sanofi Pasteur MSD, Sanofi Pasteur, Cubist, Wyeth, Merck, Pfizer, Roche, Regeneron, Jansen, Medimmune, Novavax, Novartis and GSK. IRC belongs to the Board of Directors of the CAV-AEP. AS-A has received funding from Sanofi, Pfizer and MSD for lectures and grants. ALV has received research grants from MSD and funding from Angelini Pharma for lectures and advisory boards. SKM has received funding from GSK, Pfizer, and Sanofi Pasteur for lectures and ad hoc advisory boards. The other authors have indicated that they have neither potential conflicts of interest nor financial relationships relevant to the article to disclose.

### Contributors Statement

Dr. Costanza Di Chiara and Dr. Pierre-Philippe Piché-Renaud contributed to: conceptualization, survey development, methodology, investigation, visualization, writing – original draft.

Dr. Vera Rigamonti and Dr. Anna Cantarutti contributed to: methodology, formal analysis, investigation, data curation, visualization, writing – Review & Editing.

Dr. Cristina Calvo, Dr. Irene Rivero Calle, Dr. Antoni Soriano-Arandes, Dr. Luigi Cantarutti, Dr. Daniele Donà, Dr. Andrea Lo Vecchio, Dr. Adamos Hadjipanayis, Dr. Zoi Dorothea Pana, Dr. Ioannis Kopsidas contributed to: resources, survey review and adaptation, validation, writing – Review & Editing.

Dr Adria Rose contributed to: methodology, data curation, project administration, writing – Review & Editing.

Dr. Carlo Giaquinto and Dr. Alfredo Tagarro contributed to: resources, methodology, validation, supervision, writing – Review & Editing.

Shaun K Morris contributed to: conceptualization, methodology, resources, project administration, validation, supervision, writing – Review & Editing.

All authors reviewed, edited, and approved the final version of the manuscript, authorized its submission for publication, and agree to be accountable for all aspects of the work.

### Data Availability Statement

Data are available from the corresponding author upon reasonable request.

## List of Online Resource

Online Resource 1. Survey instrument. Online Resource 2. Scores construction.

Online Resource 3. Geographical distribution of responders (N=578) by country, categorized by sortation areas in Italy, Cyprus, and Greece, and by regions in Spain.

Online Resource 4. Pediatric subspecialties of specialized participants (N=218), overall and stratified by country.

Online Resource 5. Representativeness of the restricted sample: Comparison between participants from Spain and Italy who completed the questionnaire (N=212) and those with incomplete responses (N=518).

Online Resource 6. Risk factors for severe RSV disease identified by 578 participants at the individual levels. Black bars represent correct answers, while gray bars indicate incorrect answers.

Online Resource 7. Risk factors for severe RSV disease identified by 578 participants at the population levels. Black bars represent correct answers, while gray bars indicate incorrect answers.

Online Resource 8. Perceived knowledge of previous (palivizumab) and new (nirsevimab and maternal vaccine) RSV prevention products.

Online Resource 9. Perceived effectiveness of palivizumab in preventing RSV infection outcomes.

Online Resource 10. Acceptance, attitude, and preference of respondents toward nirsevimab administration (N=578) by country.

Online Resource 11. Knowledge of RSV risk factors (Score #1) and prevention products (Score #2: palivizumab, nirsevimab, maternal RSV vaccine) among Spanish and Italian pediatricians, by likelihood to prescribe nirsevimab and the maternal vaccine.

Online Resource 12. Presence of perceived barriers to caregivers’ acceptance of nirsevimab and specific barriers identified among respondents.

Online Resource 13. Presence of perceived barriers to RSV maternal vaccine administration and specific barriers identified among respondents.

Online Resource 14. Presence of perceived barriers to RSV maternal vaccine acceptance and specific barriers identified among respondents.

Online Resource 15. Frequency with which respondents discuss influenza, COVID-19, and tetanus-diphtheria-pertussis vaccinations with pregnant women during their practice.

