## Supplementary material for "Pediatricians’ Attitudes and Knowledge of RSV Immunization Products": supplemental materials.pdf

[*]Contributed as co-first authors

***List of Online Resource***

Online Resource 1. Survey instrument.

Online Resource 2. Scores construction.

Online Resource 3. Geographical distribution of responders (N=578) by country, categorized by sortation areas in Italy, Cyprus, and Greece, and by regions in Spain.

Online Resource 4. Pediatric subspecialties of specialized participants (N=218), overall and stratified by country.

Online Resource 5. Representativeness of the restricted sample: Comparison between participants from Spain and Italy who completed the questionnaire (N=212) and those with incomplete responses (N=518).

Online Resource 6. Risk factors for severe RSV disease identified by 578 participants at the individual levels. Black bars represent correct answers, while gray bars indicate incorrect answers.

Online Resource 7. Risk factors for severe RSV disease identified by 578 participants at the population levels. Black bars represent correct answers, while gray bars indicate incorrect answers.

Online Resource 8. Perceived knowledge of previous (palivizumab) and new (nirsevimab and maternal vaccine) RSV prevention products.

Online Resource 9. Perceived effectiveness of palivizumab in preventing RSV infection outcomes.

Online Resource 10. Acceptance, attitude, and preference of respondents toward nirsevimab administration (N=578) by country.

Online Resource 11. Knowledge of RSV risk factors (Score #1) and prevention products (Score #2: palivizumab, nirsevimab, maternal RSV vaccine) among Spanish and Italian pediatricians, by likelihood to prescribe nirsevimab and the maternal vaccine.

Online Resource 12. Presence of perceived barriers to caregivers' acceptance of nirsevimab and specific barriers identified among respondents.

Online Resource 13. Presence of perceived barriers to RSV maternal vaccine administration and specific barriers identified among respondents.

Online Resource 14. Presence of perceived barriers to RSV maternal vaccine acceptance and specific barriers identified among respondents.

Online Resource 15. Frequency with which respondents discuss influenza, COVID-19, and tetanus-diphtheria-pertussis vaccinations with pregnant women during their practice.

### Respiratory Syncytial Virus (RSV) Disease and Prevention Products Survey

For the English survey, please click on the "English" language button above or change your selection by clicking on the globe logo at the top right corner of this page.Per il sondaggio in italiano, clicca sul pulsante della lingua qui sopra.Για την έρευνα στα ελληνικά κάντε κλικ στο κουμπί γλώσσας παραπάνω.Para el cuestionario en español, por favor hacer click en el botón de arriba.Respiratory Syncytial Virus (RSV) Disease and Prevention Products SurveyYou are being asked to participate in this survey as a pediatrician who may provide care to children affected by RSV. This survey will ask about Palivizumab, Nirsevimab and maternal RSV vaccination and aims to assess the current knowledge, attitudes and perceptions around RSV disease and prevention products as well as explore practitioners' insights on vaccine acceptance on RSV immunization of children and pregnant women in Canada and Europe. Survey responses on these topics may help provide insights into RSV prevention strategies and immunization products in Canada and Europe.Your participation in this study will require the completion of the following survey. This should take approximately 10 minutes of your time. The survey asks questions about your experience and knowledge of RSV, and attitudes toward RSV old and new prevention products. Your participation will be anonymous, and you will not be contacted again in the future. This survey involves minimal risk to you. The benefits, however, may impact society by helping to inform and support implementation of RSV prevention products in Canada, Europe, and elsewhere.You do not have to participate in this study if you do not want to. If you have any questions about this project, you may contact our research team at the following address: (principal investigator), (co-principal investigator); for Italy:; for Cyprus:; for Greece; and for Spain: , and. By completing this survey, you are consenting to its use in research. Once you have submitted your answers, they will be put into a database and will not be identifiable to you. This means that once you have submitted your survey, your responses cannot be withdrawn from the study. This survey was developed by the SickKids Initiative for Vaccine Acceptance and Confidence (IVAC) team and collaborators:Dr. Shaun Morris, Division of Infectious Diseases, The Hospital for Sick ChildrenDr. Pierre-Philippe Piché-Renaud, Division of Infectious Diseases, The Hospital for Sick Children Mr. Ryan Huang, Temerty Faculty of Medicine, University of TorontoDr. Upton Allen, Division of Infectious Diseases, The Hospital for Sick Children Dr. Shelly Bolotin, Public Health OntarioDr. Costanza Di Chiara, Division of Infectious Diseases, The Hospital for Sick ChildrenDr. Malini Dave, Pediatricians Alliance of OntarioDr. Tiffany Fitzpatrick, Public Health OntarioDr. Julia Orkin, Division of Pediatric Medicine, The Hospital for Sick ChildrenDr. Michelle Science, Division of Infectious Diseases, The Hospital for Sick ChildrenConsidering the anticipated impact of the study's findings on optimizing educational strategies for RSV prevention products among healthcare workers, not only in Canada but also in other countries, a collaboration has been initiated between The Hospital for Sick Children and the Penta-Child Health Research. In addition to Canada, this survey is being distributed among pediatricians working also in Italy, Spain, Cyprus, and Greece through centers affiliated with the Penta-Child Health Research Respiratory Infections Working Group.

---

Language

- ◯ English
- ◯ Italiano
- ◯ Ελληνικά
- ◯ Español

#### Information on your practice

| | |
|---|---|
| Please indicate the country of your primary practice: | ◯ Italy<br>◯ Greece<br>◯ Cyprus<br>◯ Spain |
| What is your specialty? | ◯ General Pediatrician<br>◯ Pediatrician Subspecialist<br>◯ Other {other_pediatrician_specialty} |
| What is (are) your pediatric subspecialty(ies)?<br>(Please select all that apply.) | ☐ Allergy and Immunology<br>☐ Cardiology<br>☐ Critical care (PICU)<br>☐ Dermatology<br>☐ Emergency medicine and general pediatric ward<br>☐ Endocrinology<br>☐ Gastroenterology<br>☐ Infectious Diseases<br>☐ Hematology/Oncology<br>☐ Nephrology<br>☐ Neonatology<br>☐ Neurology<br>☐ Pulmonology<br>☐ Rheumatology<br>☐ Other {other_pediatrician_subspecialty} |
| How many years have you been in practice? | ◯ Less than 5 years<br>◯ 6-10 years<br>◯ 10-25 years<br>◯ More than 25 years |
| Where did you complete your medical training? | ◯ Italy<br>◯ Greece<br>◯ Cyprus<br>◯ Spain<br>◯ Other {other_medical_training_country} |
| Which setting would you best describe your primary practice? | ◯ Urban<br>◯ Suburban<br>◯ Rural<br>◯ Remote |
| How would you describe your primary practice? | ◯ Primary Care Center<br>◯ Primary or Secondary Hospital<br>◯ Tertiary Care Academic Hospital<br>◯ Private Hospital<br>◯ Private Primary outpatient Centre<br>◯ Other {eu_other_practice_description} |

Within your primary practice, how often are you managing cases of bronchiolitis and upper respiratory tract infections during the winter season?

◯ Always ◯ Very Often ◯ Sometimes ◯ Rarely ◯ Never

What are the first two digits of your practice's postal code?

This information will be used to determine the corresponding site of participants' practice, which will help us collect geographically stratified data, such as vaccine coverage and practices that differ between geographical regions in Italy.

__________________________________

What are the first two digits of your practice?

This information will be used to determine the corresponding site of participants' practice, which will help us collect geographically stratified data, such as vaccine coverage and practices that differ between geographical regions in Greece and Cyprus.

__________________________________

What autonomous community do you practice in?

This information will be used to determine the corresponding site of participants' practice, which will help us collect geographically stratified data, such as vaccine coverage and practices that differ between geographical regions in Spain.

- ◯ - Andalusia
- ◯ - Aragon
- ◯ - Asturias
- ◯ - Balearic Islands
- ◯ - Basque Country
- ◯ - Canary Islands
- ◯ - Cantabria
- ◯ - Castile and Leòn
- ◯ - Castilla-La Mancha
- ◯ - Catalonia
- ◯ - Extremadura
- ◯ - Galicia
- ◯ - La Rioja
- ◯ - Madrid
- ◯ - Murcia
- ◯ - Navarre
- ◯ - Valencia
- ◯ - Ceuta
- ◯ - Melilla

Does your practice provide immunizations to children?

- ◯ Yes
- ◯ No

At an individual level, which of the following underlying conditions increase the risk for severe RSV disease and are thus currently recommended to receive {it_gr_palivizumab} {es_nirsevimab}? (check all that apply)

- ☐ Infants born ≤ 32 weeks gestation
- ☐ Infants 33-35 completed weeks gestation with additional risk factors
- ☐ Infants living in remote communities lacking immediate access to medical care
- ☐ Children with Trisomy 21
- ☐ Children with bronchopulmonary dysplasia/chronic lung disease (BPD/CLD)
- ☐ Children with congenital heart disease aged < 12 months
- ☐ Children with cerebral palsy
- ☐ Children with chronic renal or liver disease
- ☐ Children with severe asthma

Are you aware of national recommendations on RSV prophylaxis?
{it_rsv_awareness_national_recommendations}
{gr_rsv_awareness_national_recommendations}
{es_rsv_awareness_national_recommendations}

- ◯ Yes
- ◯ No

Are you aware of any Risk Assessment Tool for the Respiratory Syncytial Virus (RSV) Prophylaxis for High-Risk Infants?

◯ Yes
◯ No

Please, specify which Risk Assessment Tool(s)

__________________________________

At a population level, which group of infants represents the highest number of cases of severe RSV disease?

◯ Healthy infants and children under 2 years
◯ Healthy children between 2 and under 5 years
◯ Children with underlying cardiac comorbidities
◯ Children with underlying lung comorbidities
◯ Children with underlying immunodeficiencies or immunosuppression
◯ Infants/Children born prematurely
◯ Children with chronic neurologic or genetic conditions

On a scale from one to five, how would you rate your knowledge of the following RSV prevention products?

On a scale from one to five, how would you rate your knowledge of the following previous, current, and upcoming RSV prevention products?

| | 1. Very Low | 2. Below Average | 3. Average | 4. Above Average | 5. Very High |
|---|---|---|---|---|---|
| Palivizumab (Commercial name: Synagis) | ◯ | ◯ | ◯ | ◯ | ◯ |
| Nirsevimab (Commercial name: Beyfortus) | ◯ | ◯ | ◯ | ◯ | ◯ |
| Maternal vaccination during pregnancy (RSVpreF Vaccine) | ◯ | ◯ | ◯ | ◯ | ◯ |

#### Palivizumab (Commercial name: Synagis)

A monoclonal antibody against the RSV fusion protein with a half-life of 18-21 days, administered monthly during RSV season. Palivizumab has been approved for use in Europe since 1999, with proven real-world efficacy and safety in late-preterm and term infants.

A monoclonal antibody against the RSV fusion protein with a half-life of 18-21 days, administered monthly during RSV season until 2022-2023 in Spain.

In the last five years, how many of your patients have received Palivizumab?

- ◯ None
- ◯ 1-9
- ◯ 10+

{it_gr_palivizumab_administration_who} {es_palivizumab_administration_who}

- ☐ Physicians
- ☐ Nurses
- ☐ Nurse practitioners
- ☐ Trainees
- ☐ Other {palivizumab_administration_other}

For the following groups for whom Palivizumab is recommended, how important do you think Palivizumab administration is?

For the following groups for whom Palivizumab was recommended before the introduction of Nirsevimab, how important do you think the administration of Palivizumab was?

| | Very important | Important | Neutral | Not very important | Not at all important |
|---|---|---|---|---|---|
| Infants born ≤ 32 weeks gestation | ◯ | ◯ | ◯ | ◯ | ◯ |
| Infants 33-35 completed weeks gestation with additional risk factors (Risk Assessment tool score 49 to 100) | ◯ | ◯ | ◯ | ◯ | ◯ |
| Infants living in remote communities lacking immediate access to medical care | ◯ | ◯ | ◯ | ◯ | ◯ |
| Children with Trisomy 21 | ◯ | ◯ | ◯ | ◯ | ◯ |
| Children with bronchopulmonary dysplasia/chronic lung disease (BPD/CLD) | ◯ | ◯ | ◯ | ◯ | ◯ |
| Children with congenital heart disease | ◯ | ◯ | ◯ | ◯ | ◯ |

{it_gr_palivizumab_rec_proportion} {es_palivizumab_rec_proportion}

- ◯ < 50%
- ◯ 50-74%
- ◯ 75-99%
- ◯ 100%

{it_gr_palivizumab_acceptance_proportion} {es_palivizumab_acceptance_proportion}

- ◯ < 50%
- ◯ 50-74%
- ◯ 75-99%
- ◯ 100%

| | |
|---|---|
| {it_gr_palivizumab_receive_proportion} {es_palivizumab_receive_proportion} | ◯ < 50%<br>◯ 50-74%<br>◯ 75-99%<br>◯ 100% |
| {it_gr_palivizumab_additional_groups} {es_palivizumab_additional_groups} | ◯ Yes {palivizumab_addgroups_specify}<br>◯ No |

How effective do you think Palivizumab is against the following consequences of RSV infection?

How effective did you think Palivizumab was against the following consequences of RSV infection?

| | Very effective | Somewhat effective | Ineffective |
|---|---|---|---|
| Medically attended outpatient visit | ◯ | ◯ | ◯ |
| Emergency room visit | ◯ | ◯ | ◯ |
| Hospital Admission | ◯ | ◯ | ◯ |
| ICU Admission | ◯ | ◯ | ◯ |
| Death | ◯ | ◯ | ◯ |

| | |
|---|---|
| {it_gr_palivizumab_barriers_administration} {es_palivizumab_barriers_administration} | ◯ Yes<br>◯ No |

Please check all that apply:

☐ Low priority for the product
☐ Perception of poor effectiveness by pediatricians
☐ Process to order the product
☐ Cost to the system
☐ Parental hesitancy
☐ Challenges in identifying patients that are eligible
☐ Other {palivizumab_barriers_administration_other}

| | |
|---|---|
| {it_gr_palivizumab_barriers} {es_palivizumab_barriers} | ◯ Yes<br>◯ No |

Please check all that apply:

☐ Lack of awareness of the product
☐ Perception of poor effectiveness of the product
☐ Process to access the product
☐ Multiple doses requiring multiple visits
☐ Cost to the family
☐ Concerns for potential adverse effects {palivizumab_barriers_adverse_effects}
☐ Other {palivizumab_barriers_other}

#### Nirsevimab

A monoclonal antibody against the RSV fusion protein that has an extended half-life allowing for a single dose throughout an RSV season. It has been shown to protect healthy preterm infants from RSV-associated lower respiratory tract infection and was approved for use in Europe in November 2022.

Once Nirsevimab becomes available in [country], which would be your preferred setting and timing for administration?

◯ During birth hospitalization
◯ During the first well-baby visit
◯ At a specific vaccine clinic
◯ Other {nirsevimab_administration_preferred_other}

Which is your preferred setting and timing for the administration of Nirsevimab?

◯ During birth hospitalization
◯ At Primary Care Centres
◯ During the first well-baby visit
◯ At a specific vaccine clinic
◯ Other {es_nirsevimab_administration_preferred_other}

Assuming a similar cost of the product and similar side-effect profile, once Nirsevimab become available in [country], how likely are you to recommend it over Palivizumab?

◯ Very likely ◯ Likely ◯ Neutral ◯ Unlikely ◯ Very unlikely

{it_gr_nirsevimab_administration_likelihood} {es_nirsevimab_administration_likelihood}

◯ Very likely ◯ Likely ◯ Neutral ◯ Unlikely ◯ Very unlikely

**Amongst the following product characteristics, how important for you are each in making your recommendation to parents about Nirsevimab?**

| | Very important | Important | Neutral | Not very important | Not at all important |
|---|---|---|---|---|---|
| Safety in clinical trial | ○ | ○ | ○ | ○ | ○ |
| Efficacy in clinical trial | ○ | ○ | ○ | ○ | ○ |
| Real-world effectiveness | ○ | ○ | ○ | ○ | ○ |
| ECDC recommendation | ○ | ○ | ○ | ○ | ○ |
| Italian pediatric society and Italian pediatric infectious diseases society (SITIP) | ○ | ○ | ○ | ○ | ○ |
| Ped Society position statement | ○ | ○ | ○ | ○ | ○ |
| Spanish Ministry of Health recommendations | ○ | ○ | ○ | ○ | ○ |
| Asociación Española de Pediatría Sociedad Española de Infectología Pediátrica (AEP SEIP) position statement | ○ | ○ | ○ | ○ | ○ |
| Expert opinion | ○ | ○ | ○ | ○ | ○ |
| Ease of administration and access | ○ | ○ | ○ | ○ | ○ |

Are there any other product characteristics that are important for you when making recommendations to parents about Nirsevimab?

○ Yes {important_characteristic_recommendation_other}
○ No

{it_gr_nirsevimab_use_group} {es_nirsevimab_use_group}

All infants in the first year of life {nirsevimab_use_all_infant_first_y}
Only high-risk infants {nirsevimab_use_high_risk_infants}

Please specify (check all that apply)

☐ Preterm birth
☐ BDP/CLD
☐ Congenital heart disorder
☐ Trisomy 21
☐ Immunocompromised
☐ Infants lacking access to immediate medical care
☐ Other {nirsevimab_use_other_high_risk_groups}

{it_nirsevimab_seasonal_text}
{gr_nirsevimab_seasonal_text}
{es_nirsevimab_seasonal_text}

○ all-year
○ seasonal

{it_gr_nirsevimab_acceptance_barriers}
{es_nirsevimab_acceptance_barriers}

○ Yes
○ No

Please specify (check all that apply)

☐ Replacing a well-established product with a new product
☐ Lack of safety and efficacy data
☐ Lack of real-world data
☐ Location of administration
☐ Giving a healthy baby a monoclonal antibody (mAb) product
☐ Lack of awareness
☐ Concerns for potential adverse effects {nirsevimab_barriers_adverse_effects}
☐ Other {nirsevimab_barriers_other}

{it_gr_nirsevimab_administration_barriers}
{es_nirsevimab_administration_barriers}

◯ Yes
◯ No

Please specify (check all that apply)

☐ Introduction of a new product
☐ Lack of safety and efficacy data
☐ Lack of real-world data
☐ Use in groups not traditionally recommended in Palivizumab recommendations
☐ Storage
☐ Availability of healthcare providers to administer vaccines
☐ Patient access to healthcare
☐ Potential for emerging resistance
☐ Other {nirsevimab_administration_barriers_other}

#### Maternal RSV Vaccination

Maternal RSV vaccinations are given as a single injection late in pregnancy to initiate the development of antibodies that pass through the placenta. The pre-F subunit maternal vaccine, RSVpreF, was approved by the EMA in September 2023 for use in pregnancy between 32 weeks and 36 weeks of gestation. However, national recommendations have not yet been released.

Once both Nirsevimab and recommendations on the maternal RSV vaccine become available in [country], how likely are you to recommend the RSV maternal vaccine over Nirsevimab?

◯ Very likely ◯ Likely ◯ Neutral ◯ Not likely ◯ Very unlikely

Once recommendations for the maternal RSV vaccine become available in [country], do you think acceptance for maternal RSV vaccination will be higher than Nirsevimab?

◯ Much higher ◯ Higher ◯ No difference ◯ Lower ◯ Much lower

**Amongst the following product characteristics, how important for you are each in recommending the maternal RSV vaccine, once recommendations become available?**

| | Very important | Important | Neutral | Not very important | Not at all important |
|---|---|---|---|---|---|
| Safety in clinical trial | ◯ | ◯ | ◯ | ◯ | ◯ |
| Efficacy in clinical trial | ◯ | ◯ | ◯ | ◯ | ◯ |
| Real-world effectiveness | ◯ | ◯ | ◯ | ◯ | ◯ |
| ECDC recommendation | ◯ | ◯ | ◯ | ◯ | ◯ |
| SIP SITIP position statement | ◯ | ◯ | ◯ | ◯ | ◯ |
| Ped Society position statement | ◯ | ◯ | ◯ | ◯ | ◯ |
| Spanish Ministry of Health recommendations | ◯ | ◯ | ◯ | ◯ | ◯ |
| AEP SEIP position statement | ◯ | ◯ | ◯ | ◯ | ◯ |
| Expert opinion | ◯ | ◯ | ◯ | ◯ | ◯ |
| Ease of administration and access | ◯ | ◯ | ◯ | ◯ | ◯ |

Are there any other product characteristics that will be important for you when making recommendations to parents about maternal RSV vaccine?

◯ Yes

rtant_characteristic_recommendation_other_maternal}

◯ No

Do you anticipate any barriers to RSV vaccine administration during pregnancy in your patient community?

◯ Yes
◯ No

Please specify (check all that apply):

☐ Lower prioritization of vaccine due to competing demands
☐ Introducing vaccine in pregnancy
☐ Administration of multiple vaccines during pregnancy
☐ Lack of safety and efficacy data
☐ Lack of real-world data
☐ Storage
☐ Availability of healthcare providers to administer vaccines
☐ Concerns for potential adverse effects {barriers_rsv_administration_adv_effects}
☐ Patient access to healthcare
☐ Cost
☐ Potential for emerging resistance
☐ Other {barriers_rsv_administration_specify}

Do you anticipate any barriers to RSV vaccine acceptance during pregnancy?

◯ Yes
◯ No

Please specify (check all that apply):

☐ Lack of safety and efficacy data
☐ Lack of real-world data
☐ Lack of awareness
☐ Concerns for potential adverse effects {barriers_rsv_vac_acceptance_adv_effects}
☐ Concerns for potential adverse effects to infant {barriers_rsv_vac_acceptance_adv_effects_infant}
☐ Other {barriers_rsv_vaccine_acceptance_specify}

**How often do you discuss each of the following vaccinations in pregnancy in your practice (even if you are not the one giving it)?**

| | Always | Very Often | Sometimes | Rarely | Never |
|---|---|---|---|---|---|
| Influenza | ◯ | ◯ | ◯ | ◯ | ◯ |
| COVID-19 | ◯ | ◯ | ◯ | ◯ | ◯ |
| Tetanus-diphtheria-pertussis (Tdap) | ◯ | ◯ | ◯ | ◯ | ◯ |

If influenza, COVID-19, and RSV vaccines in pregnancy become available in [country], how likely are you to recommend all these vaccines as a respiratory prevention approach for mothers and babies?

◯ Always ◯ Very Often ◯ Sometimes ◯ Rarely ◯ Never

#### Infant RSV Vaccination

Should an infant RSV vaccine become available in [country], how likely are you to recommend it over passive immunization with Nirsevimab?

◯ Very likely ◯ Likely ◯ Neutral ◯ Not likely ◯ Very unlikely

Should both the maternal and infant RSV vaccines become recommended and available in [country], how likely are you to recommend the infant vaccine over the maternal vaccine to your patients?

◯ Very likely ◯ Likely ◯ Neutral ◯ Not likely ◯ Very unlikely

Should the infant RSV vaccine become available in [country], how likely do you think patients will show a greater acceptance toward the infant vaccine compared to the maternal vaccine?

◯ Very likely ◯ Likely ◯ Neutral ◯ Not likely ◯ Very unlikely

Please provide any other comments on RSV disease and prevention products ______________________________

***Methods, Online Resource 2.***

Scores construction.

We developed three distinct scores to quantify (1) knowledge of RSV risk factors and (2) knowledge of RSV prevention products among participants. Higher scores indicate a greater level of knowledge.

Key questions were selected for each score to capture the most critical aspects of these areas.

Score #1: Knowledge of RSV risk factors

The questions *"At an individual level, which of the following underlying conditions increase the risk for severe RSV disease and are thus currently recommended to receive palivizumab/nirsevimab?"* (with *"palivizumab"* for Italy and *"nirsevimab"* for Spain) and *"At a population level, which group represents the largest proportion of severe RSV disease?"* were considered. The first question included nine evaluations, of which five were correct, and four were incorrect. The maximum possible score for this question was 9, with one point deduced for each incorrect classification. For the second question, one point was awarded if the response was correct. The first and second scores were summed, resulting in a total score #1 ranging from 5 to 10.

Score #2: Knowledge of RSV prevention products

This score was based on the question, *"On a scale from one ("very low") to five ("very high"), how would you rate your knowledge of the following previous, current, and upcoming RSV prevention products?"*. The score was determined by asking participants to rate their knowledge of palivizumab, nirsevimab, and maternal vaccination on a scale of 1 (very low) to 5 (very high). Each product was assigned a score of 1-5. These three scores constituted the total score, ranging from a minimum of 3 to a maximum of 15.

The raw scores were standardized on a scale of 0 to 10. Subsequently, these normalized scores were categorized into three groups: 'low,' 'medium,' and 'high.' The cut-off points for these categories were determined by the 25th and 75th percentiles of the distribution of normalized scores. Given the study's primary objective of identifying pediatricians with lower levels of knowledge, the 'low' category was of particular.

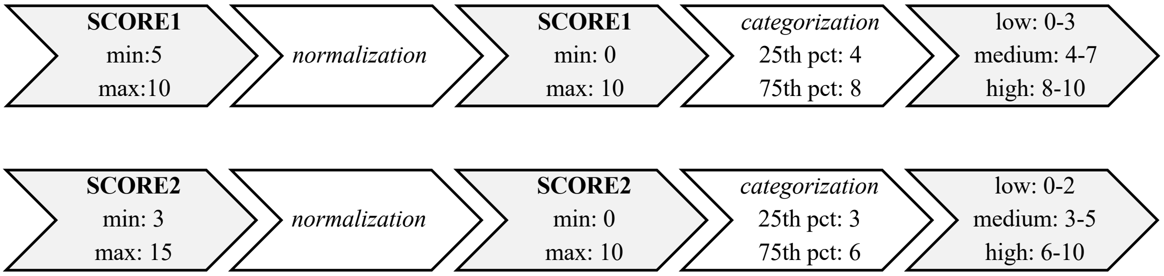
SCORE1
min:5
max:10
normalization
SCORE1
min: 0
max: 10
categorization
25th pct: 4
75th pct: 8
low: 0-3
medium: 4-7
high: 8-10
SCORE2
min: 3
max: 15
normalization
SCORE2
min: 0
max: 10
categorization
25th pct: 3
75th pct: 6
low: 0-2
medium: 3-5
high: 6-10

***Figure, Online Resource 3***. Geographical distribution of responders (N=578) by country, categorized by sortation areas in Italy, Cyprus, and Greece, and by regions in Spain.

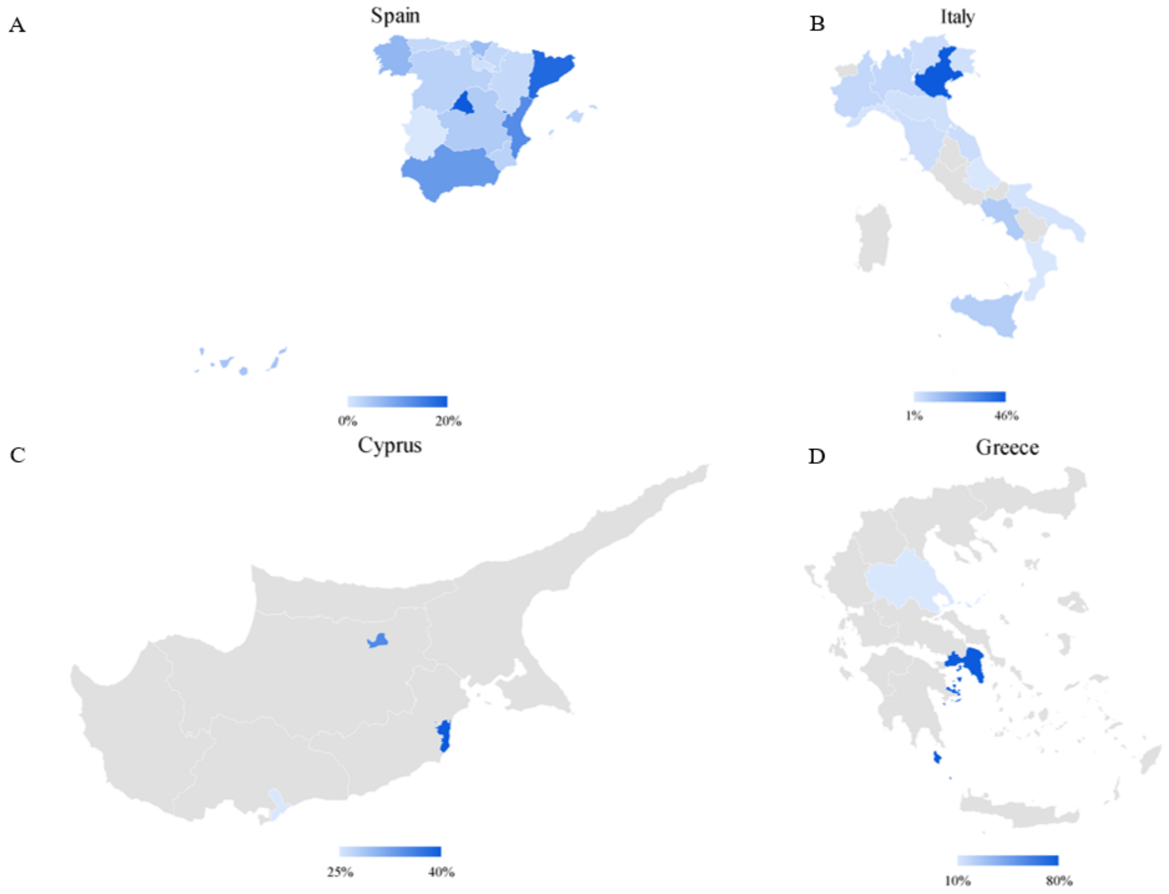

***Table, Online Resource 4.*** Pediatric subspecialties of specialized participants (N=218), overall and stratified by country.

| Subspecialty N (%) | Overall (N=218) | Spain (N=108) | Italy (N=97) | Cyprus (N=3) | Greece (N=10) |
|---|---|---|---|---|---|
| **Allergy and immunology** | 6 (2.8%) | 1 (0.9%) | 5 (5.2%) | 0 | 0 |
| **Cardiology** | 7 (3.2%) | 4 (3.7%) | 3 (3.1%) | 0 | 0 |
| **Critical care** | 16 (7.3%) | 10 (9.3%) | 5 (5.2%) | 0 | 1 (10%) |
| **Dermatology** | 1 (0.5%) | 0 | 1 (1%) | 0 | 0 |
| **Emergency medicine and general pediatric ward** | 31 (14.2%) | 19 (17.6%) | 12 (12.4%) | 0 | 0 |
| **Endocrinology** | 11 (5.1%) | 6 (5.6%) | 5 (5.2%) | 0 | 0 |
| **Gastroenterology** | 13 (6%) | 7 (6.5%) | 6 (6.2%) | 0 | 0 |
| **Infectious diseases** | 60 (27.5%) | 21 (19.4%) | 32 (33%) | 1 (33.4%) | 6 (60%) |
| **Haematology/oncology** | 14 (6.4%) | 7 (6.5%) | 6 (6.2%) | 0 | 1 (10%) |
| **Nephrology** | 12 (5.5%) | 7 (6.5%) | 5 (5.2%) | 0 | 0 |
| **Neonatology** | 31 (14.2%) | 18 (16.7%) | 10 (10.3%) | 0 | 3 (30%) |
| **Neurology** | 4 (1.8%) | 2 (1.9%) | 2 (2.1%) | 0 | 0 |
| **Pulmonology** | 9 (4.1%) | 5 (4.6%) | 3 (3.1%) | 1 (33.3%) | 0 |
| **Rheumatology** | 9 (4.1%) | 4 (3.7%) | 5 (5.2%) | 0 | 0 |
| **Other** | 13 (6%) | 12 (11.1%) | 1 (1%) | 0 | 0 |

Percentages do not sum to 100% as responses were not mutually exclusive.

***Table, Online Resource 5.*** Representativeness of the restricted sample: Comparison between participants from Spain and Italy who completed the questionnaire (N=212) and those with incomplete responses (N=518).

| | Sample | | |
|---|---|---|---|
| | **N=518** | **N=212** | **SMD*** |
| **Country** | | | 0.1024 |
| **Spain** | 290 (56.0%) | 108 (50.9%) | |
| **Italy** | 228 (44.0%) | 104 (49.1%) | |
| **Specialty** | | | 0.1557 |
| **Pediatricians subspecialist** | 205 (39.9%) | 101 (47.6%) | |
| **General pediatrician** | 309 (60.1%) | 111 (52.4%) | |
| **Years in practice** | | | 0.0613 |
| **<=10** | 145 (28.3%) | 66 (31.1%) | |
| **>10** | 367 (71.7%) | 146 (68.9%) | |
| **Description of primary practice** | | | |
| **Primary care&Primary outpatient center** | 275 (53.3%) | 104 (49.1%) | 0.0841 |
| **Primary/secondary care hospital&Private hospital** | 82 (15.9%) | 32 (15.1%) | 0.0221 |
| **Academic hospital&Tertiary care hospital** | 159 (30.8%) | 76 (35.8%) | 0.1062 |
| **Manage of RSV cases** | | | 0.0275 |
| **Always&Very often** | 480 (92.7%) | 198 (93.4%) | |
| **Sometimes&Rarely&Never** | 38 (7.3%) | 14 (6.6%) | |
| **Provision of immunizations to children** | | | 0.0411 |
| **Yes** | 443 (85.9%) | 185 (87.3%) | |
| **No** | 73 (14.1%) | 27 (12.7%) | |

*Standardized Mean Difference.

***Figure, Online Resource 6.*** Risk factors for severe RSV disease identified by 578 participants at the individual levels. Black bars represent correct answers, while gray bars indicate incorrect answers.

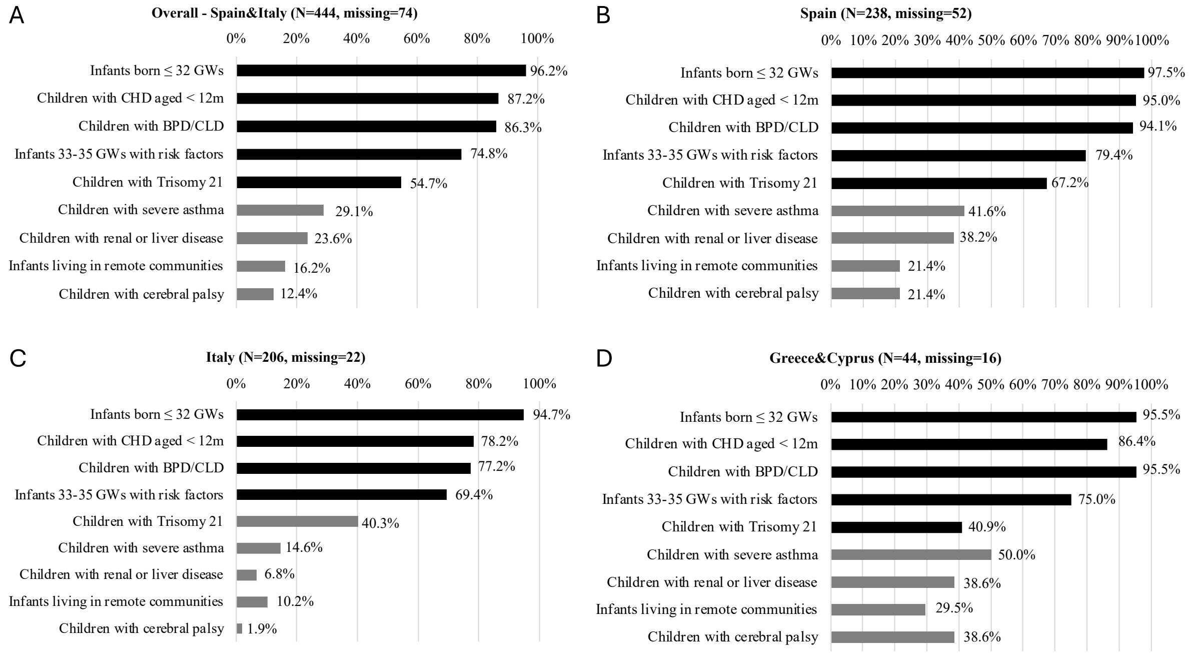


CHD= congenital heart disease; BPD/CLD= bronchopulmonary dysplasia/chronic lung disease.

***Figure, Online Resource 7.*** Risk factors for severe RSV disease identified by 578 participants at the population levels. Black bars represent correct answers, while gray bars indicate incorrect answers.

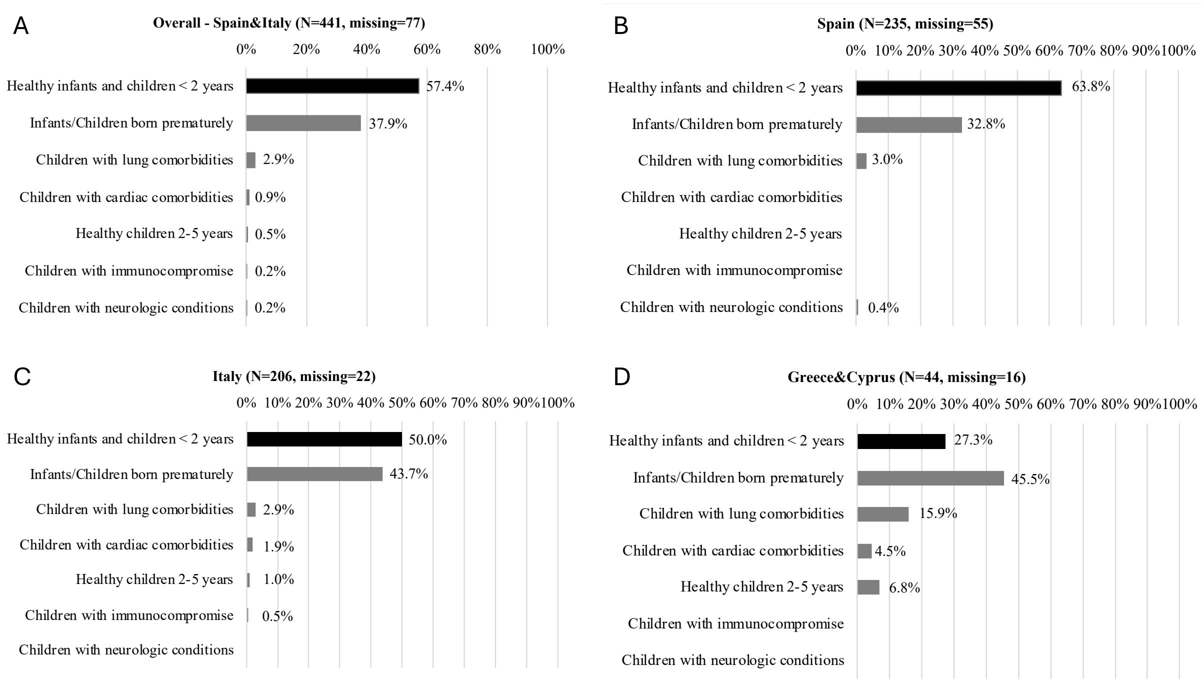

***Figure, Online Resource 8.*** Perceived knowledge of previous (palivizumab) and new (nirsevimab and maternal vaccine) RSV prevention products.

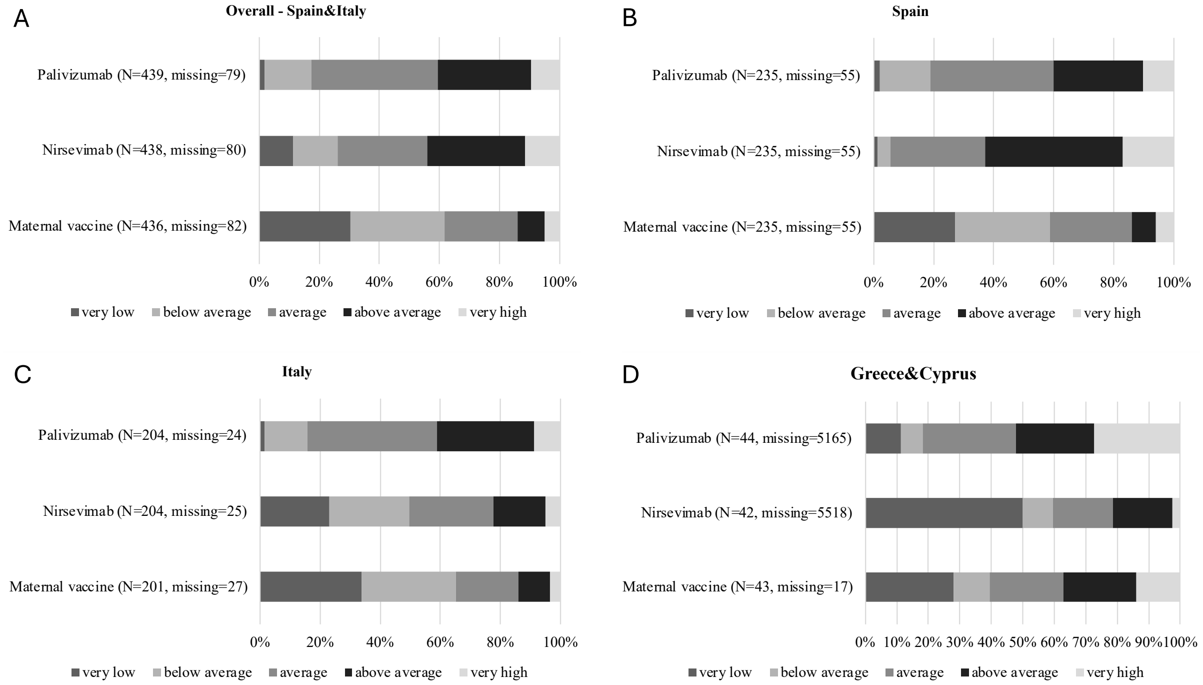

***Figure, Online Resource 9.*** Perceived effectiveness of palivizumab in preventing RSV infection outcomes.

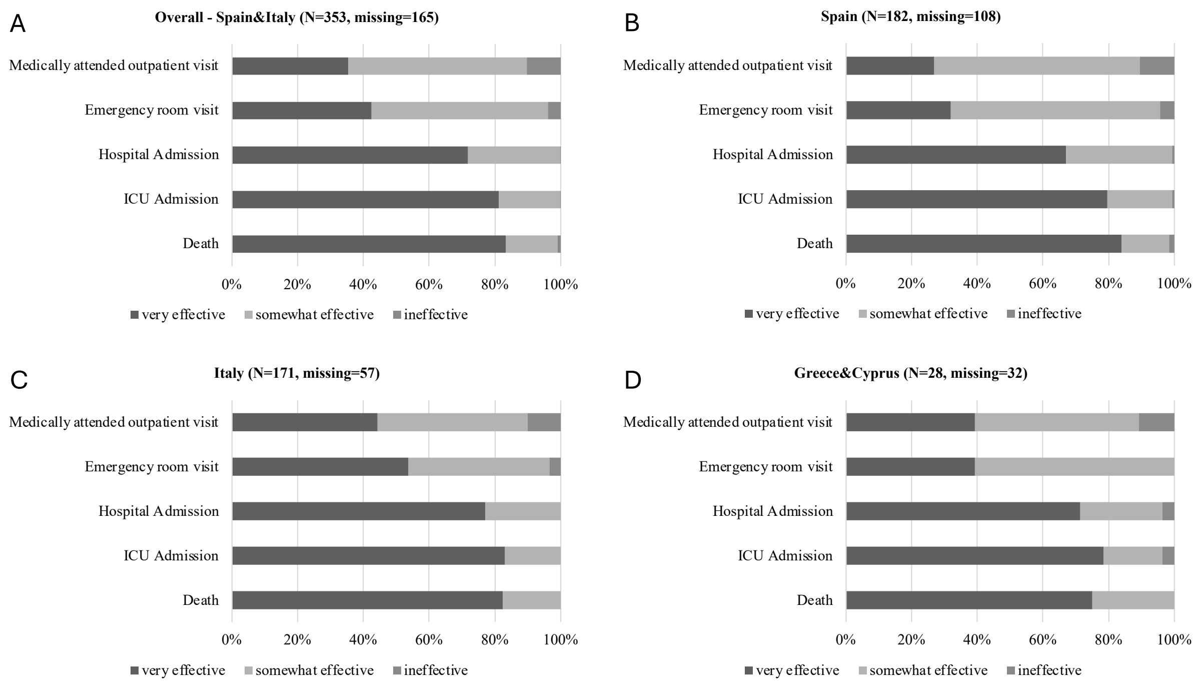


ICU=intensive care unit.

***Table, Online Resource 10.*** Acceptance, attitude, and preference of respondents toward nirsevimab administration (N=578) by country.

| **Administration's preference** | **Spain (N=290)** | **Italy (N=228)** | **Greece&Cyprus (N=60)** |
|---|---|---|---|
| **Likelihood of administering nirsevimab** | *(missing=110)* | *(missing=46)* | *(missing=27)* |
| Very likely | 173 (96.1%) | 138 (75.8%) | 15 (45.5%) |
| Likely | 4 (2.2%) | 35 (19.2%) | 14 (42.4%) |
| Neutral | 0 (0.0%) | 9 (4.9%) | 2 (6.1%) |
| Unlikely | 1 (0.6%) | 0 (0.0%) | 1 (3%) |
| Very unlikely | 2 (1.1%) | 0 (0.0%) | 1 (3%) |
| **Target population** | | | |
| All infants | 171 (96.1%) *(missing=112)* | 148 (88.1%) *(missing=60)* | 10 (33.3%) *(missing=30)* |
| Only high-risk infants | 18 (11.8%) *(missing=138)* | 33 (21.2%) *(missing=72)* | 24 (72.7%) *(missing=27)* |
| ***Only high-risk infants*** | *(n/18)* | *(n/33)* | *(n/24)* |
| Preterm birth | 18 (100.0%) | 33 (100.0%) | 23 (95.8%) |
| BDP/CLD | 17 (94.4%) | 33 (100.0%) | 22 (91.7%) |
| CHD | 18 (100.0%) | 31 (93.9%) | 20 (83.3%) |
| Trisomy 21 | 18 (100.0%) | 22 (66.7%) | 16 (66.7%) |
| Immunocompromised | 18 (100.0%) | 28 (84.9%) | 18 (75.0%) |
| Infants lacking access to immediate medical care | 8 (44.4%) | 18 (54.6%) | 8 (33.3%) |
| Other | 1 (5.6%) | 1 (3.0%) | 0 (0.0%) |
| **Preferred setting** | *(missing=108)* | *(missing=46)* | *(missing=27)* |
| During birth hospitalization | 164 (90.1%) | 128 (70.3%) | 14 (42.4%) |
| During the first well-baby visit | 4 (2.2%) | 29 (15.9%) | 12 (36.4%) |
| At a specific vaccine clinic | 0 (0.0%) | 17 (9.3%) | 6 (18.2%) |
| At primary care centers | 14 (7.7%) | 0 (0.0%) | 0 (0.0%) |
| Other | 0 (0.0%) | 8 (4.4%) | 1 (3%) |
| **Preferred time** | *(missing=112)* | *(missing=54)* | *(missing=27)* |
| Any time of the year | 47 (26.4%) | 71 (40.8%) | 8 (24.2%) |
| Seasonal | 131 (73.6%) | 103 (592%) | 25 (75.8%) |

***Table, Online Resource 11***. Knowledge of RSV risk factors (Score #1) and prevention products (Score #2: palivizumab, nirsevimab, maternal RSV vaccine) among Spanish and Italian pediatricians, by likelihood to prescribe nirsevimab and the maternal vaccine.

| | **Knowledge of RSV risk factors (Score #1)** | | |
|---|---|---|---|
| | **Low (N=34)** | **Medium (N=93)** | **High (N=85)** |
| **Pediatricians' likelihood of prescribing nirsevimab** | | | |
| Likely | 33 (97.1%) | 90 (96.8%) | 82 (96.5%) |
| Neutral/unlikely | 1 (2.9%) | 3 (3.2%) | 3 (3.5%) |
| **Pediatricians' likelihood of prescribing the RSV maternal vaccine** | | | |
| Likely | 25 (73.5%) | 69 (74.2%) | 63 (74.1%) |
| Neutral/unlikely | 9 (26.5%) | 24 (25.8%) | 22 (25.9%) |

| | **Knowledge of RSV prevention products (Score #2)** | | |
|---|---|---|---|
| | **Low (N=18)** | **Medium (N=118)** | **High (N=76)** |
| **Pediatricians' likelihood of prescribing nirsevimab** | | | |
| Likely | 17 (94.4%) | 114 (96.6%) 4 | 74 (97.4%) |
| Neutral/unlikely | 1 (5.6%) | (3.4%) | 2 (2.6%) |
| **Pediatricians' likelihood of prescribing the RSV maternal vaccine** | | | |
| Likely | 14 (77.8%) | 90 (76.3%) | 53 (69.7%) |
| Neutral/unlikely | 4 (22.2%) | 28 (23.7%) | 23 (30.3%) |

***Figure, Online Resource 12***. Presence of perceived barriers to caregivers' acceptance of nirsevimab and specific barriers identified among respondents.

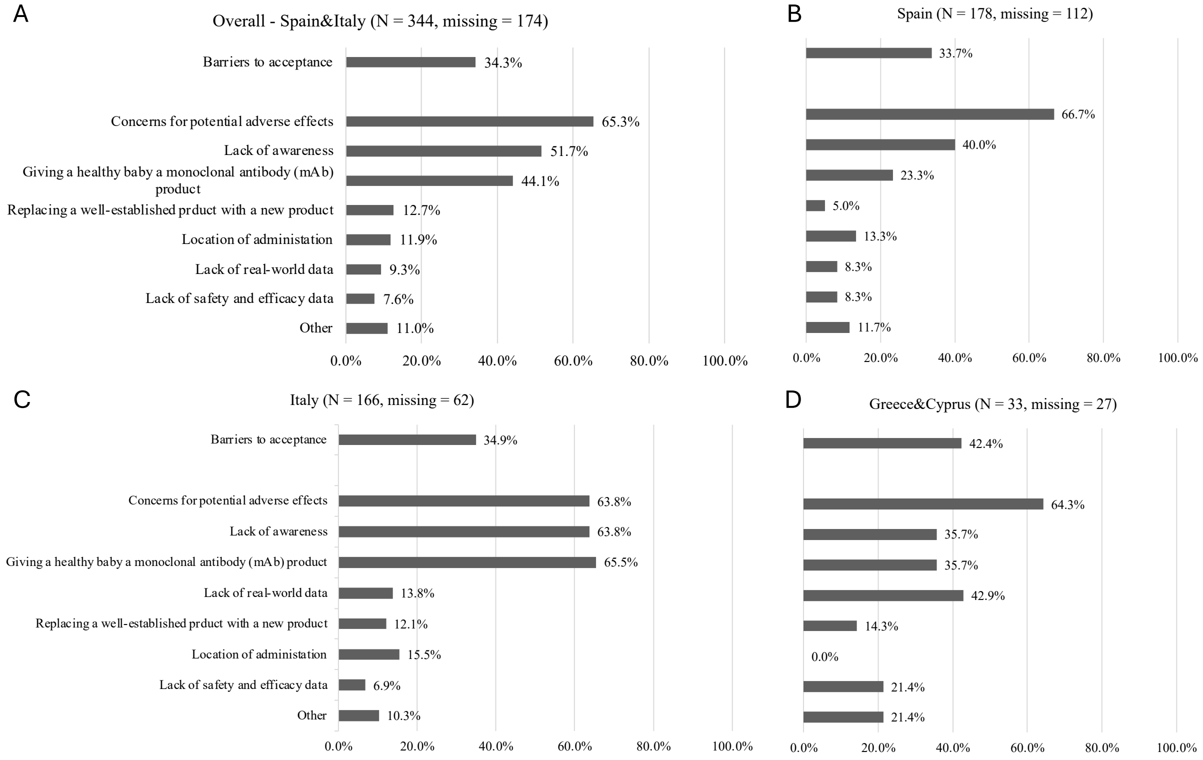

***Figure, Online Resource 13***. Presence of perceived barriers to RSV maternal vaccine administration and specific barriers identified among respondents.

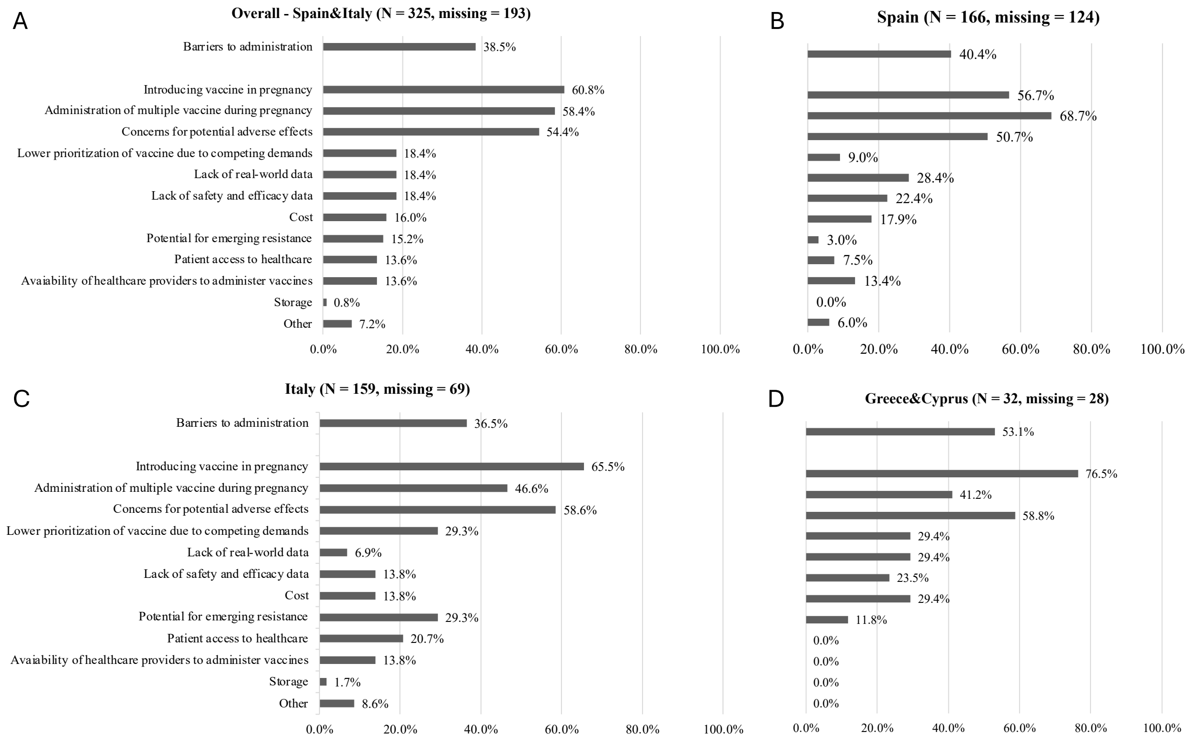

***Figure, Online Resource 14***. Presence of perceived barriers to RSV maternal vaccine acceptance and specific barriers identified among respondents.

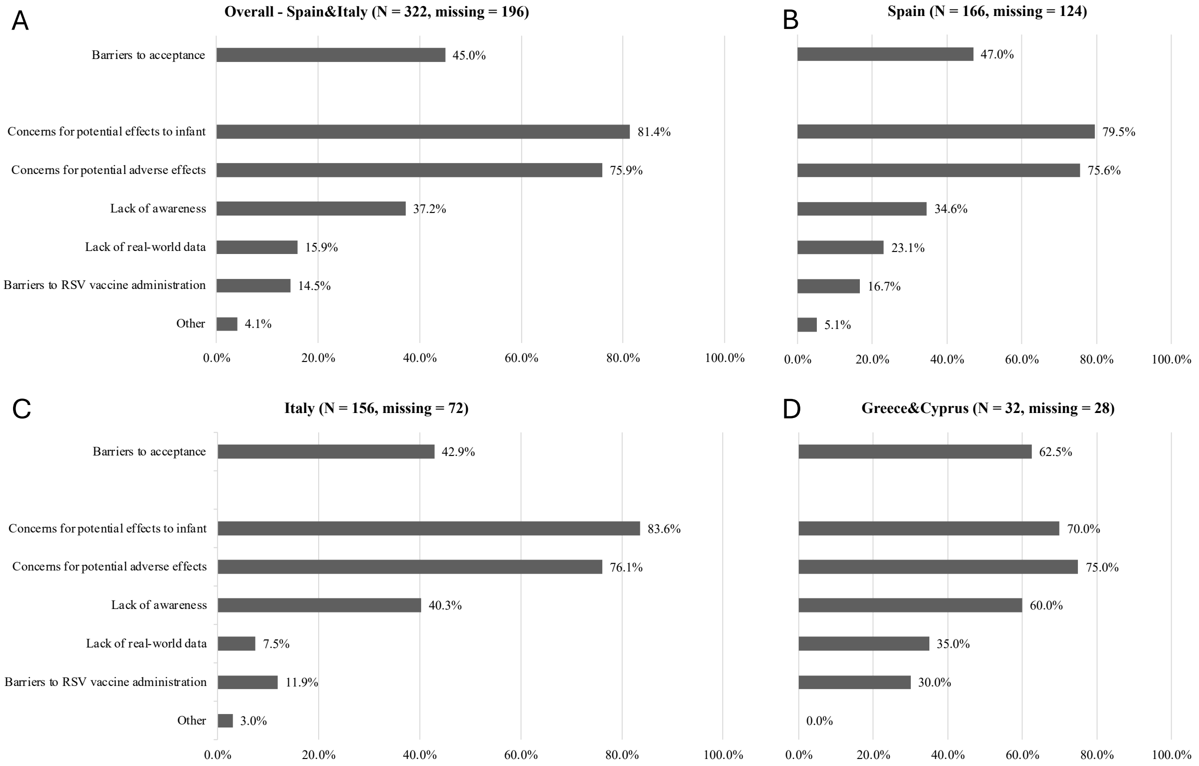

*Figure, Online Resource 15*. Frequency with which respondents discuss influenza, COVID-19, and tetanus-diphtheria-pertussis vaccinations with pregnant women during their practice.

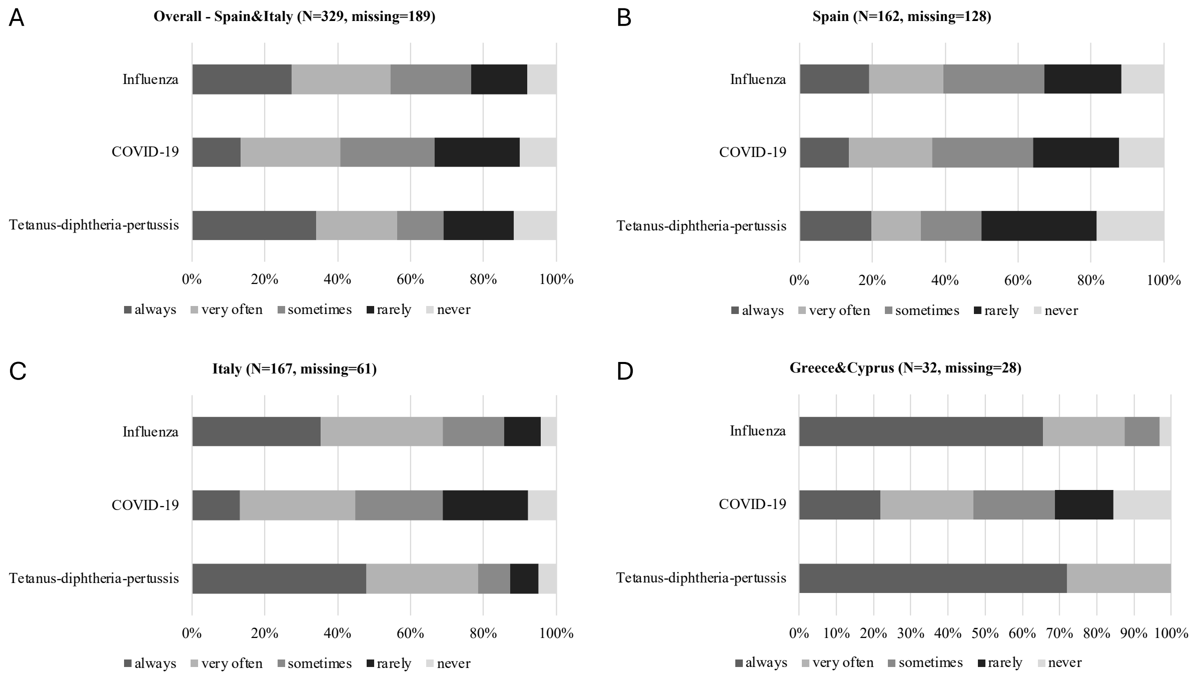
